# Genomic and eco-epidemiological investigations in Uruguay reveal local Chikungunya virus transmission dynamics during its expansion across the Americas in 2023

**DOI:** 10.1101/2023.08.17.23294156

**Authors:** Analía Burgueño, Marta Giovanetti, Vagner Fonseca, Noelia Morel, Mauricio Lima, Emerson Castro, Natália R. Guimarães, Felipe C. M. Iani, Victoria Bormida, Maria Noel Cortinas, Viviana Ramas, Leticia Coppola, Ana I. Bento, Leticia Franco, Jairo Mendez Rico, José Lourenço, Luiz Carlos Junior Alcantara, Hector Chiparelli

## Abstract

Uruguay experienced its first Chikungunya virus outbreak in 2023, resulting in a significant burden to its healthcare system. We conducted analysis based on real-time genomic surveillance (30 novel whole genomes) to offer timely insights into recent local transmission dynamics and eco-epidemiological factors behind its emergence and spread in the country.

## Text

Chikungunya virus (CHIKV) is an emerging arbovirus posing significant public health challenges in the Americas. Uruguay reported its first cases in late April to early May 2023, with 51 autochthonous and 19 imported cases, raising concerns about sustained transmission (1). This event is part of a larger trend in the Americas, where CHIKV was first detected in 2013 on the island of Saint Martin. The Americas have experienced over a million cases in the first year after CHIKV introduction, and by early 2023 over 214,000 cases had been reported (2).

Climate change is a contributing factor to CHIKV’s spread, with higher temperatures and altered rainfall patterns creating new habitats for its main vector -*Aedes* mosquitoes (3). Unplanned urbanization may also drive its spread, since *Aedes* mosquitoes prefer urban environments, mainly breeding in water containers within households (4). Uruguay’s ecological conditions and interconnectedness with neighboring countries endemic to CHIKV further influence local transmission potential (4). Understanding the combination of factors driving transmission is crucial for effective surveillance, prevention, and control.

Despite efforts, significant gaps remain in understanding the genomic diversity and evolution of circulating CHIKV strains in Uruguay. To address this, we partnered with Montevideo’s Central Public Health Laboratory for genomics and modeling studies. These efforts yielded valuable insights into viral dynamics, aiding the development of effective future control and surveillance strategies.

Residual clinical samples were collected from CHIKV-positive patients (Ct ≤35) between February and May 2023. Nanopore technology, was used to generate the first Uruguay’s 30 whole genome sequences (Accession numbers: OR360566-OR360595). Using this data, we constructed phylogenetic trees to explore the evolutionary and epidemiological links between CHIKV in Uruguay to those of other sequences of this viral genotype sampled globally. Detailed methodologies are available in the **Supplementary Material**.

Additionally, time series data for confirmed, suspected, and probable infections were downloaded from the PAHO website (5). A measure of climate-driven transmission potential (index P) was estimated as in (6), using satellite climate data (7) and human incubation and infectious periods as in (8).

Reported cases presented a seasonal signal associated with the local 2022-2023 summer period (**Figure 1A**). The correlation between estimated climate-driven transmission potential and reported cases was high at 0.93, pointing to the role of natural climatic variation in the small CHIKV outbreak. Across Uruguay, transmission potential was estimated to be highest in the Northwest departments bordering Argentina, and lowest in the South including departments surrounding the capital Montevideo (**Figure 1B**). These observations strongly suggest that local climate is modulating the emergence of CHIKV in Uruguay, which provides spatio-temporal opportunities for control and mitigation planning.

**Figure 1.**
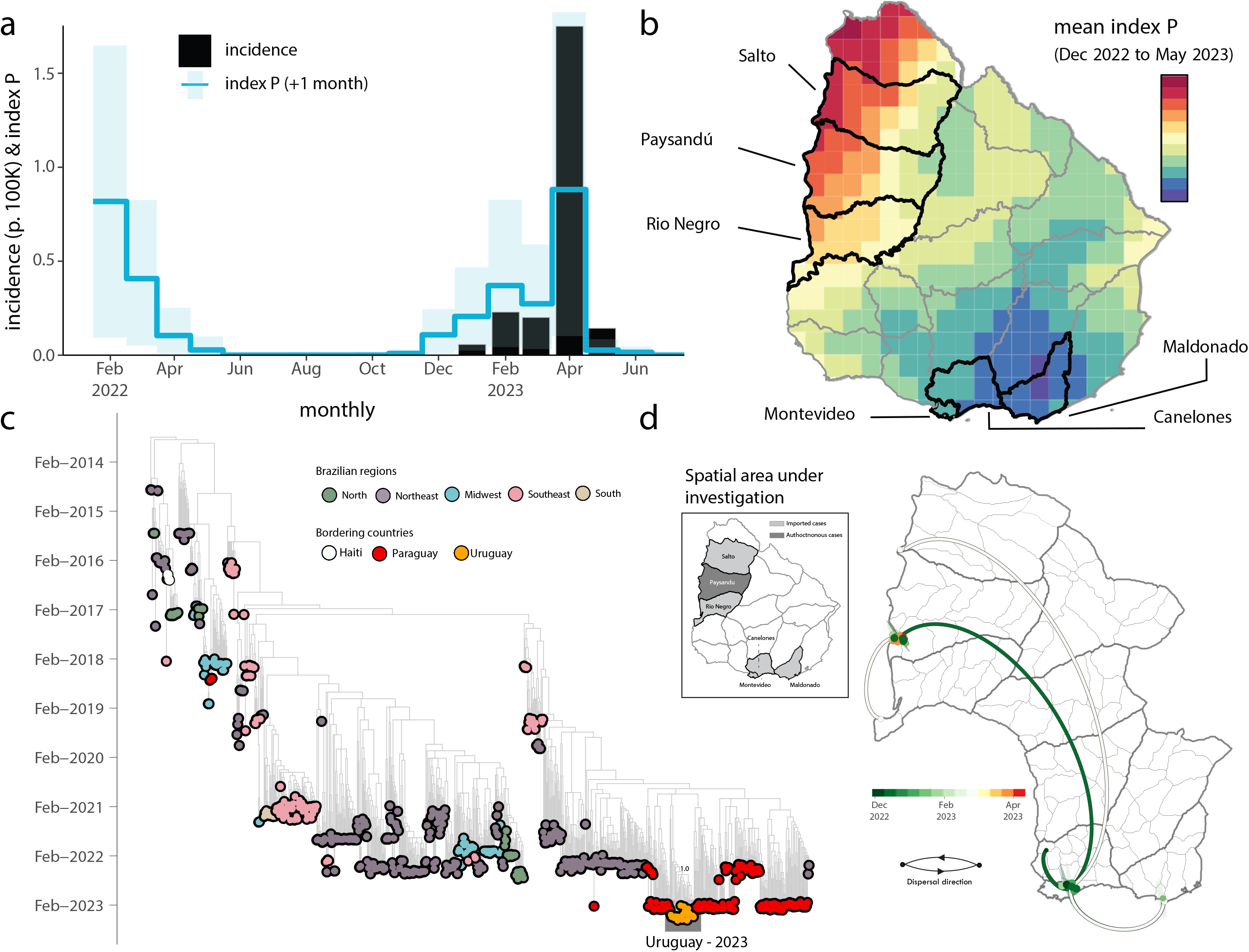
Eco-epidemiological assessment and temporal trend of CHIKV transmission in Uruguay. **a)** Time series of reported CHIKV cases (black, bars) and estimated climate-driven transmission potential (index P, blue, lines show the mean and shaded area the 95% quantile). The index P is shifted +1 month into the future (the index tends to precede cases, see (5). **b)** Map with mean index P at resolution available in climate data. Official departments are shown, highlighted (black and named) are those for which new genomes were generated. **c)** Time-resolved maximum likelihood tree including the newly complete genome sequence from Uruguay (n=30) generated in this study combined with publicly available sequences (n=819) from GenBank collected up to July 20th, 2023. Colors indicate geographic location of sampling. Support for branching structure is shown by bootstrap values at key nodes.; **d)** Spatiotemporal reconstruction of the spread of CHIKV-ECSA in Uruguay. Circles represent nodes of the maximum clade credibility phylogeny, colored according to their inferred time of occurrence (scale shown). Shaded areas represent the 80% highest posterior density interval and depict the uncertainty of the phylogeographic estimates for each node. Solid curved lines denote the links between nodes and the directionality of movement. Curved lines denote the direction of transmission in the anti-clockwise direction.

Out of the tested positive samples, 30 showed sufficient DNA (≥ 2 ng/μL) for library preparation. The average cycle threshold (Ct) values for PCR were 23.50, ranging from 18 to 32 (**Table S1**). During the sequencing process, an average coverage of 97% was achieved, with a range of 84% to 99.5%. The genotyping assignment confirmed that all samples belonged to the East/Central/South African (ECSA) lineage. Samples were obtained from six different departments (**Table S1 and Figure 1)**. Median age was 45 years, ranging from 10 to 84 years. Approximately 70% (n=21) were female (**Table S1**). A total of 22 were identified as autochthonous cases, while the remaining 8 reported a travel history to Brazil (n=1) and Paraguay (n=7).

A phylodynamic investigation was performed including the 30 newly generated CHIKV genomes in addition to 819 representative ECSA referenced strains (**Fig. 1c**). This analysis revealed that the novel genomes formed a well-structured monophyletic clade, indicating a close genetic relationship. While genetic data alone cannot establish the directionality of transmission, our findings suggest a probable origin in Paraguay (**Fig. 1c**). However, it is crucial to consider the limited availability of whole genome sequences from other Uruguayan departments and neighboring countries like Argentina. This scarcity of sequences restricts our ability to better describe and understand the molecular epidemiology of viral strains at a regional level.

Focusing solely on the novel genomes, we reconstructed viral movements across different departments in Uruguay. The analysis indicated that the mean time of origin for these samples was in mid-December 2022, with a 95% highest posterior density (HPD) ranging from early-December 2022 to early-January 2023. Notably, viruses from this clade spread multiple times from the South Departments (Canelones, Montevideo, and Maldonado) towards the Midwest (Salto, Paysandú, and Rio Negro) (**Figure 1d**).

## Conclusion

The surge in autochthonous Chikungunya cases in Uruguay and in the Americas more generally raises concerns for public health. Climate change, unplanned urbanization and the connectedness within bordering countries that are endemic will have likely contributed to emergence in Uruguay. Swift and comprehensive action is imperative to control mosquito populations and prevent further transmission. Learning from past experiences and enhancing regional cooperation are crucial steps in effectively combating these vector-borne diseases and safeguarding public health. There is a pressing need to intensify sequencing efforts across the different South American countries to enhance resolution of phylogenetics in order to better understand the ongoing source-sync dynamics of CHIKV during its emergence in the continent.

## Data Availability

Accession numbers: OR360566 to OR360595

## Ethics statement

This project was reviewed and approved by the Pan American Health Organization Ethics Review Committee (PAHOERC) (Ref. No. PAHO-2016-08-0029). The samples used in this study were de-identified residual samples from the routine diagnosis of arboviruses in the Paraguayan public health laboratory, which is part of the public network within the Uruguayan Ministry of Health.

## Biographical sketch

Dr. Analía Burgueño is a distinguished researcher at the Emerging Virus Laboratory, located within the Laboratorio Central de Salud Pública in Montevideo, Uruguay. Her primary focus revolves around molecular diagnosis and sequencing of various critical pathogens, including SARS-CoV-2, arboviruses such as DENV, ZIKV, CHIKV, YFV, and other pathogens that significantly impact Public Health.

## Acknowledgments

This study was supported by the National Institutes of Health USA grant U01 AI151698 for the United World Arbovirus Research Network (UWARN) and the CRP-ICGEB RESEARCH GRANT 2020 Project CRP/BRA20-03, Contract CRP/20/03. M. Giovanetti’s funding is provided by PON “Ricerca e Innovazione ‘‘2014-2020. The authors would also like to acknowledge the Global Consortium to Identify and Control Epidemics – CLIMADE, (T.O., L.C.J.A., E.C.H., J.L., M.G.) (https://climade.health/).

## Author contributions

Conception and design: M.G., V.F., J.L., L.C.J.A. and H.C.; Investigations: A.B., M.G., V.G., M.N.M., M.L., E.C., N.R.G., V.B., M.N.C., V.R., L.C., A.I.B., L.F., J.M.R., J.L., L.C.J.A., H.C.; Data Analysis: M.G., V.F., and J.L; Visualization: M.G., V.F., and J.L.; Writing – Original: M.G., and J.L.; Revision: A.B., M.G., V.G., M.N.M., M.L., E.C., N.R.G., V.B., M.N.C., V.R., L.C., A.I.B., L.F., J.M.R., J.L., L.C.J.A., H.C.; Resources: L.F., J.M.R., L.C.J.A., H.C .

## References

1. Pan-American Health Organization. Chikungunya cases in the Americas. 2023. https://www.paho.org/en/news/4-5-2023-rising-cases-experts-discuss-chikungunya-spread-americas

2. Giovanetti M, Vazquez C, Lima M, Castro E, Rojas A, Gomez de la Fuente A, et al. Rapid epidemic expansion of chikungunya virus East/Central/South African lineage, Paraguay. Emerg Infect Dis. 2023; 16;15(2), 23–24.

3. Tjaden, N.B., Suk, J.E., Fischer, D. et al. Modelling the effects of global climate change on Chikungunya transmission in the 21st century. Sci Rep. 2017; 7,(9), 38–93.

4. Weaver SC. Urbanization and geographic expansion of zoonotic arboviral diseases: mechanisms and potential strategies for prevention. Trends Microbiol. 2013; 21(8), 360–3.

5. Pan-American Health Organization, CHIKV Weekly Report. PAHO. 2023. https://www3.paho.org/data/index.php/en/mnu-topics/chikv-en/550-chikv-weekly-en.html

6. Nakase, T., Giovanetti, M., Obolski, U. et al. Global transmission suitability maps for dengue virus transmitted by Aedes aegypti from 1981 to 2019. Sci Data 10, 275 (2023). https://doi.org/10.1038/s41597-023-02170-7

7. Hersbach, H., Muñoz Sabater, J., Nicolas Rozum, I., Simmons Vamborg, F. A., Bell, B., Berrisford, P., Biavati, G., Buontempo, C., Horányi, A. J., Peubey, C., Radu, R., Schepers, D., Soci, C., Dee, D., Thépaut, J-N. (2018): Essential climate variables for assessment of climate variability from 1979 to present. Copernicus Climate Change Service (C3S).

8. Pereira Gusmão Maia Z, Mota Pereira F, do Carmo Said RF, Fonseca V, Gräf T, de Bruycker Nogueira F, Brandão Nardy V, Xavier J, Lima Maia M, Abreu AL, Campelo de Albuquerque CF, Kleber Oliveira W, Croda J, de Filippis AMB, Venancio Cunha R, Lourenço J, de Oliveira T, Faria NR, Junior Alcantara LC, Giovanetti M. Return of the founder Chikungunya virus to its place of introduction into Brazil is revealed by genomic characterization of exanthematic disease cases. Emerg Microbes Infect. 2019 Dec 27;9(1):53–57. doi: 10.1080/22221751.2019.1701954. PMID: 31880218; PMCID: PMC6968431.

