## Supplementary_Appendix for "Genomic and eco-epidemiological investigations in Uruguay reveal local Chikungunya virus transmission dynamics during its expansion across the Americas in 2023"

### Escalating Chikungunya in the Americas: tracing the trail of the epidemic in Uruguay.

### Supplementary Material

#### Details on Methods

##### Sample collection and whole genome sequencing

Residual clinical samples, collected between February and May 2023, were obtained from patients who tested positive for CHIKV using molecular screening at the Laboratorio Central de Salud Publica of Uruguay, in Montevideo. Only genomes with Ct values <35 were selected for sequencing.

Extracted RNA was first converted to cDNA using the SuperScript IV Reverse Transcriptase kit (Invitrogen) and subjected to sequencing multiplex PCR (35 cycles) as previously described (1). DNA library preparation was conducted using the Ligation Sequencing kit (Oxford Nanopore Technologies) and Native Barcoding Expansion 1-96 kit (Oxford Nanopore Technologies), following the reaction conditions as previously described in (2). Sequencing was performed for up to 24 hours on a MinION device and consensus sequences were obtained using the Genome Detective software (3).

The arbovirus genotyping tool was used to investigate sequence genotypes (4). To investigate CHIKV evolution in Uruguay, the newly generated complete genome sequences were combined with globally available sequences retrieved from NCBI up to July 20th, 2023. We excluded sequences without sampling date and location as well as sequences that covered less than 50% of the virus genome.

All sequences were aligned using MAFFT (5) and manually edited to remove artifacts using Aliview (6). The GTR nucleotide substitution model, which was inferred as the best-fit model by the ModelFinder application implemented in IQ-TREE2 (7), was used to estimate maximum Likelihood (ML) phylogenetic trees. The tree topology's robustness was determined using 1,000 bootstrap replicates. TempEst (8) was used to assess the presence of a temporal signal, and the BEAST package (9) was used to infer time-scaled phylogenetic trees. To estimate the most appropriate molecular clock model for the Bayesian phylogenetic analysis, we used a stringent model selection analysis that included both path-sampling (PS) and steppingstone (SS) procedures (10). For all datasets, the uncorrelated relaxed molecular clock model was chosen by estimating marginal likelihoods using the codon-based SRD06 model of nucleotide substitution and the nonparametric Bayesian Skyline coalescent model. To model the phylogenetic diffusion of detected community transmission clades we used a flexible relaxed random walk diffusion model (11, 12) that accommodates branch-specific variation in rates of dispersal with a Cauchy distribution and a jitter window site of 0.01 (13, 14). Latitude and longitude coordinates were assigned to each sequence. BEAST v1.10.4 was used for the MCMC analyses, which were run in duplicate for 100 million interactions and sampled every 10,000 steps in the chain. Tracer was used to assess convergence for each run (effective sample size for all relevant model parameters >200). After discarding the initial 10% as burn-in, MCC trees for each run were summarized using TreeAnnotator. Finally, we extracted and mapped spatiotemporal information embedded in the posterior trees using the R package 'seraphim' version 1.0 (14).

Epidemiological data

Epidemiological data of weekly notified and laboratory confirmed cases CHIKV in Uruguay were obtained and curated from the PAHO data repository for Chikungunya (15). Confirmed infections are defined as “a suspected or probable chikungunya case with a chikungunya test with positive result” (as stated on the PAHO platform).

**References**

1. Giovanetti M, Vazquez C, Lima M, Castro E, Rojas A, Gomez de la Fuente A, et al. Rapid epidemic expansion of chikungunya virus East/Central/South African lineage, Paraguay. Emerg Infect Dis. 2023; 16;15(2), 23-24.

2. Quick J, Grubaugh ND, Pullan ST, Claro IM, Smith AD, Gangavarapu K, et al. Multiplex PCR method for MinION and Illumina sequencing of Zika and other virus genomes directly from clinical samples. Nat Protoc. 2017;12(12), 61–76.

3. Vilsker M, Moosa Y, Nooij S, Fonseca V, Ghysens Y, Dumon K, et al. Genome Detective: an automated system for virus identification from high-throughput sequencing data. Bioinformatics. 2019 Mar 1;35(5), 871–3

4. Fonseca V, Libin PJK, Theys K, Faria NR, Nunes MRT, Restovic MI, et al. A computational method for the identification of dengue, Zika and chikungunya virus species and genotypes. PLoS Negl Trop Dis. 2019;13(1), 7-31.

5. Katoh K, Rozewicki J, Yamada KD. MAFFT online service: multiple sequence alignment, interactive sequence choice and visualization. Brief Bioinformatics. 2019;20(4), 1160-1166.

6. Larsson A. AliView: a fast and lightweight alignment viewer and editor for large datasets. Bioinformatics. 2014;30(3), 276–8.

7. Nguyen L-T, Schmidt HA, von Haeseler A, Minh BQ. IQ-TREE: a fast and effective stochastic algorithm for estimating maximum-likelihood phylogenies. Mol Biol Evol. 2015;32(1), 268–74.

8. Rambaut A, Lam TT, Max Carvalho L, Pybus OG. Exploring the temporal structure of heterochronous sequences using TempEst (formerly Path-O-Gen). Virus Evol. 2016;2(1), 1-7.

9. Suchard MA, Lemey P, Baele G, Ayres DL, Drummond AJ, Rambaut A. Bayesian phylogenetic and phylodynamic data integration using BEAST 1.10. Virus Evol. 2018;4(2), 14-16.

10. Baele, G., Li, W. L., Drummond, A. J., Suchard, M. A. & Lemey, P. Accurate model selection of relaxed molecular clocks in bayesian phylogenetics. Mol. Biol. Evol. 2013;30(1), 239–243.

11. Lemey P, Rambaut A, Welch JJ, Suchard MA. 2010. Phylogeography takes a relaxed random walk-in continuous space and time. Mol Biol Evol. 2010;27(3), 1877–1885.

12. Pybus OG, Suchard MA, Lemey P, Bernardin FJ, Rambaut A, Crawford FW, Gray RR, Arinaminpathy N, Stramer SL, Busch MP, Delwart EL. Unifying the spatial epidemiology and molecular evolution of emerging epidemics. Proc Natl Acad Sci USA. 2012;2(109), 15066–15071.

13. Dellicour, S. et al. Relax, keep walking - a practical guide to continuous phylogeographic inference with BEAST. Mol. Biol. Evol. 2021; 38,(9), 3486–3493.

14. Dellicour, S., Rose, R., Faria, N. R., Lemey, P. & Pybus, O. G. SERAPHIM:studying environmental rasters and phylogenetically informed movements. Bioinformatics. 2016; 32(9), 3204–3206.

15.Pan-American Health Organization, CHIKV Weekly Report. PAHO. 2023. https://www3.paho.org/data/index.php/en/mnu-topics/chikv-en/550-chikv-weekly-en.html
